# A Flexible Data-Driven Framework for COVID-19 Case Forecasting Deployed in a Developing-world Public Health Setting

**DOI:** 10.1101/2021.11.01.21260020

**Authors:** Sansiddh Jain, Avtansh Tiwari, Nayana Bannur, Ayush Deva, Siddhant Shingi, Vishwa Shah, Mihir Kulkarni, Namrata Deka, Keshav Ramaswami, Vasudha Khare, Harsh Maheshwari, Soma Dhavala, Jithin Sreedharan, Jerome White, Srujana Merugu, Alpan Raval

## Abstract

Forecasting infection case counts and estimating accurate epidemiological parameters are critical components of managing the response to a pandemic. This paper describes a modular, extensible framework for a COVID-19 forecasting system, primarily deployed during the first Covid wave in Mumbai and Jharkhand, India. We employ a variant of the SEIR compartmental model motivated by the nature of the available data and operational constraints. We estimate best fit parameters using Sequential Model-Based Optimization (SMBO), and describe the use of a novel, fast and approximate Bayesian model averaging method (ABMA) for parameter uncertainty estimation that compares well with a more rigorous Markov Chain Monte Carlo (MCMC) approach in practice. We address on-the-ground deployment challenges such as spikes in the reported input data using a novel weighted smoothing method. We describe extensive empirical analyses to evaluate the accuracy of our method on ground truth as well as against other state-of-the-art approaches. Finally, we outline deployment lessons and describe how inferred model parameters were used by government partners to interpret the state of the epidemic and how model forecasts were used to estimate staffing and planning needs essential for addressing COVID-19 hospital burden.

**CCS CONCEPTS:**

- **Applied computing** → **Health care information systems**; Forecasting;
- **Computing methodologies** → *Modeling methodologies*.

## 1 INTRODUCTION

The ongoing COVID-19 pandemic has spurred intense interest in epidemiological forecasting models. The need for robust solutions has been especially pressing in dense populations across the developing world, with their limited health resource availability and long lead times for addressing shortfalls.

To reduce mortality, it is important to ensure adequate capacity availability of critical health care resources. There is a need for forecasting reported infections at a local level to inform capacity planning, model the effects of policy changes and prepare for potential scenarios. In this paper, we describe a deployed forecasting framework that was used in Mumbai, India, a densely populated city, as well as in other resource-constrained regions such as the state of Jharkhand, India, during the first Covid infection wave. Partners for data and usage of the solution were the Brihanmumbai Municipal Corporation and the Integrated Disease Surveillance Programme, respectively, at the two locations.

### 1.1 Operational Challenges

Successful deployment of epidemiological models requires addressing operational challenges inherent in the public health landscape.

#### Data limitations

Data collection and management activities during a pandemic are highly impacted due to severe demands on public health authorities, leading to data discrepancies. In the face of these data challenges, it is important to choose model families whose complexity matches that of the available data.

#### Need for rich insights and what-if-scenarios

The choice and implementation of correct policy is often predicated not just on expected case counts but also on changes in underlying epidemiological parameters. It is therefore important to construct models that are highly interpretable with independently verifiable parameters.

#### Dynamic user requirements

Forecasting needs are highly dynamic as the epidemic progresses. In the early stages, when containment is critical, accurate regional forecasts for short horizons are valuable. In later stages, the emphasis shifts to estimation of hospital burden and to uncertainty around those projections. A robust yet flexible modeling framework that can support multiple application-specific needs is thus essential.

These challenges motivate the need for a forecasting framework that can support data-driven parameter estimation and uncertainty quantification for interpretable models. Such models cannot be directly learned using gradient-based learning approaches. Further, variations in data semantics and user requirements necessitate a solution approach that is agnostic to the choice of model class and application-specific prediction quality criteria.

### 1.2 Epidemiological Forecasting Problem

The general epidemiological model parameter estimation problem can be stated as follows. For a given spatial region, let *X* [*t*] denote a multi-variate time series of case counts corresponding to different demographics groups and disease stages. Let *M*_*θ*_ be a black-box epidemiological forecasting model parameterized by *θ* ∈ Θ ⊆ ℝ^k^ that generates a future forecast from historical observations. Given a time series loss function ℒ, the objective is to determine the optimal parameters *θ* ^*^ such that the loss between the predicted and observed data over a specified horizon [*t*_*i*_, *t* _*j*_] is minimized:

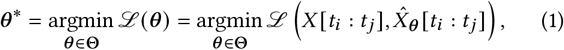

where 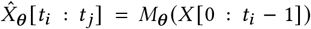. To estimate parameter uncertainty, it is also necessary to generate the resulting forecast distribution over the time horizon.

#### Deployment scenario

In our deployment scenario, the primary data comprised counts of *confirmed, active, recovered*, and *deceased* cases. For certain regions, severity-stratified counts and testing data were also available. Reliable counts of in-flow and out-flow cases and within-region mobility indicators were rarely available. While public health partner requirements changed over time, they were largely focused on hospital burden estimation over a horizon of 30–45 days. The main need was to ensure adequate medical resources while minimizing excess capacity that would remain under-utilized. A readily interpretable relative accuracy criterion such as Mean Absolute Percentage Error (MAPE) in primary case count forecasts was well-aligned with our partner needs, but the relative importance of the four different case counts could vary.

### 1.3 Main Contributions

We built a multi-purpose epidemiological forecasting framework with public health requirements in mind. This system was used to drive decision-making and planning in Mumbai and Jharkhand, India during the first Covid wave. Our framework consumes aggregate case count data culled by government officials from health facilities, and outputs burden estimates that were used by relevant authorities for subsequent planning of personnel and supplies. This paper discusses the main elements of our forecasting framework and makes the following contributions.

- *System and Process Design*. [Section 3]. We describe a practical, modular, and extensible learning-based epidemic case forecasting system that is customizable to individual regions and application scenarios. The system consists of modules for data ingestion, preprocessing and exploratory analysis, model fitting, scenario-conditioned forecasting, and application-specific report generation.
- *Modeling Methodology*. [Sections 4 and 5]. We present techniques for model and loss-agnostic estimation of parameters via sequential model based optimization (SMBO) [7]. Combining SMBO sampling with Bayesian model averaging enables fast approximate quantification of forecast uncertainty. We demonstrate this is empirically comparable to a more rigorous Markov Chain Monte Carlo (MCMC) approach but computationally faster. We develop smoothing methods to handle data issues arising out of reporting delays.
- *Epidemiological Model Choice*. [Section 4]. We argue that conditions of interpretability and identifiability warrant a simple variant of the SEIR compartmental model especially when only confirmed, active, recovered and deceased case counts are observed.
- *Empirical Results*. [Section 5]. We present extensive empirical analyses detailing the optimization of relevant hyperparameters, field predictive performance for the city of Mumbai, and comparison with other state-of-the-art models hosted by ReichLab [1].
- *Deployment Lessons*. [Section 6]. We summarize the key lessons from deployment of our modeling framework in Mumbai and Jharkhand. The audience for this work consists of applied researchers working on practical forecasting and public health officials.

## 2 RELATED WORK

The work presented here builds on four areas of epidemiological modeling: a) forecasting, b) parameter estimation (with uncertainty), c) modeling with data limitations, and d) public health deployment.

### Epidemiological Forecasting

The COVID-19 pandemic and associated global forecasting challenges [1, 24] have spurred new research on modeling infectious disease spread. There are three broad classes of models. (a) *Compartmental models* assume that individuals in a population at any given time are assigned to one of several states, called compartments, and move between these compartments. Examples include the SIR [25] and SEIR [22] models. Over the last year, variants of the SEIR model have been widely used to study the COVID-19 pandemic project hospital burden [28, 36, 37], incorporating aspects such as age-stratification [2, 20], asymptomatic transmission [34], and effects of social distancing measures [8, 16]. (b) *Agent-based models* [17, 38] simulate interactions and disease stage transitions of individual agents. (c) *Curve-fitting models* fit parameterized curves to data. Examples include the exponential growth model and the IHME CurveFit model [21].

### Model Parameters and Uncertainty Estimation

Multiple works address the problem of epidemiological parameter estimation with forecast uncertainty [9, 11, 12, 15], but these techniques are usually specific to the model class and involve assumptions on the generative process. Recently, there has been increased focus on the practical aspects of model fitting such as choice of training duration [30] and identifiability issues [27, 32]. Model-agnostic evaluation of forecasts is another related area of interest [1, 19, 35]. While our deployment was based on compartmental models, the proposed techniques are model-agnostic and can be adapted to any application-specific loss function.

### Modeling with Data Limitations

Epidemiological modeling in the developing world is plagued with data paucity and quality issues. Various studies have focused on understanding transmission dynamics in such limited data settings [4, 26, 33]. In our current work, we discuss some of these challenges and possible resolution via appropriate model choices and data preprocessing.

### Public Health Deployment

Practical use of epidemiological models in public health response requires a holistic view of government priorities, policy levers, and processes. Work in economic epidemiology [10, 31] is targeted towards supporting decision-making related to interventions and policy choices. Multiple organizations [1, 29] currently share automated COVID case forecasts with relevant public health authorities, but the forecasts are not always customized for decision-making. Our deployment involved a two-way partnership with the government, providing precise capacity planning guidance and insights on the pandemic dynamics per requirements.

## 3 SOLUTION FRAMEWORK

This section provides an overview of our end-to-end epidemic forecasting system, as visualized in Figure 1.

**Figure 1:**
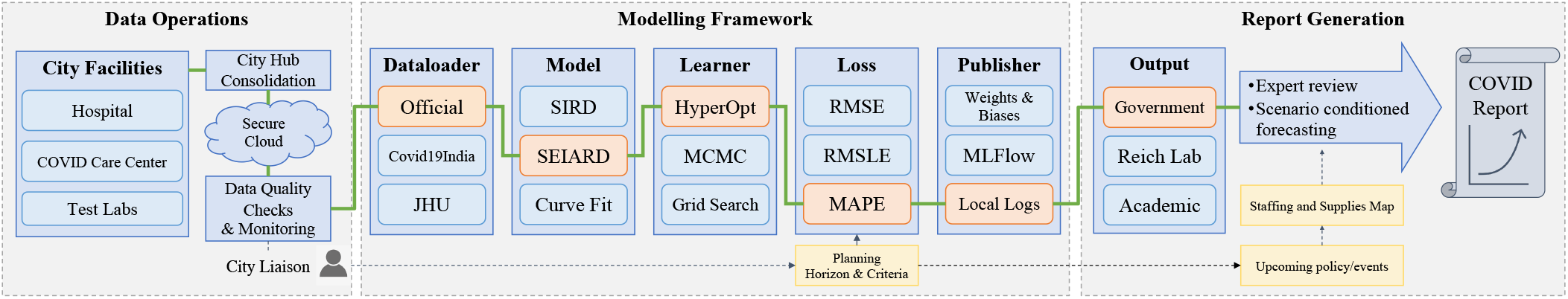
COVID forecasting framework. The three-phase pipeline consists of a) *data operations*: working with government officials to consolidate and store data collected from various sources throughout the city, b) a modular *modelling framework* for fitting and forecasting, and c) *report generation* of customized forecasts for clients. The specific components for deployment with government partners are highlighted in orange and connected by a green line delineating the workflow.

### 3.1 Data Operations

Partnership with government and official data access was enabled in the early days of the pandemic by providing proof-of-concept forecasts that initially used case counts from public sources such as Covid19India [13] and JHU [18].

Data schema for aggregate case counts was defined to facilitate automated forecasting. However, realisation of this schema across upstream providers proved to be a challenge. Issues faced included those described in the Introduction: duplicate records, point-of-entry data entry errors, inconsistencies, and errors in reporting transitions between care levels and disease stages. We therefore developed point-of-entry validation, along with partial automation of data consolidation and de-duplication. This semi-automated process was aided by sharing spreadsheets with partners on our cloud storage. Our pipeline performed quality checks and anonymized sensitive data before exposing it to the modelling team via an SQL-compliant database.^1^ We also maintained descriptive data visualizations to monitor abnormal patterns. This pipeline was later replicated across partners.

### 3.2 Modelling Framework

In order to flexibly support multiple forms of case count data, epidemiological models, and loss functions, our modelling framework has a modular design with five high-level components: data loaders, models, learners, loss functions, and publishers,^2^ each suited to multivariate time series forecasting. This extensible and scalable framework allows rapid experimentation with different modelling techniques and data sources of varying complexity, as well as a platform for comparing results. Examples of these components are shown in Figure 1. Modelling details are discussed in Section 4.

### 3.3 Report Generation

The content and format of forecast presentations varied depending on the audience. The solution framework allowed for model outputs to be consumed in a variety of ways: as time-series forecasts, planning reports (for public authorities) or data pushes (for submissions to ReichLab (Section 5.5)).

Forecasts were either routine (for monthly planning) or *ad hoc* (to address rapidly evolving scenarios). Forecast requests, compiled by team members physically located in COVID ‘war rooms’, included information on recent changes in mobility, expected changes in policy over the planning horizon and notes on data – to appropriately define plausible parameter ranges. Requests were used to generate forecasts and organized into deciles of the forecast distribution (anchored on a future date determined by the planning horizon). Specific scenarios (mapped to deciles of the forecast distribution) were selected to represent an appropriate ‘planning scenario’, a ‘low case scenario’ and a ‘high case scenario’. These interactions not only allowed public authorities to understand the state of the pandemic through changes in model parameters, but also led to the consideration of information unavailable to the model (expected changes to testing strategy, treatment policy, and non-pharmaceutical interventions such as shelter-in-place orders).

In the capacity computation module, projected active case numbers and trends in utilization (occupancy rates and length of stay) were used to estimate facility-level demand for personnel, quarantine beds, oxygen-supported beds and intensive care units. These estimates were distilled into final recommendations consisting of projected cases for a 30–45 day horizon, potential shortfalls, and recommended increases to capacity, based on resource capacity proportions shared by government partners (see Figure 12).

## 4 MODELING METHODOLOGY

This section describes our approach to parameter estimation and uncertainty quantification. We also discuss smoothing strategies for handling data spikes and the choice of epidemiological model used in the COVID-19 engagement.

### 4.1 Parameter Fitting with SMBO

Given the nonlinear nature of the optimisation problem of Equation 1, we employ a Sequential Model-Based Optimization (SMBO) method to explore the searchspace of parameters. We use the Expected Improvement (EI) criterion [23] to return the best parameter set at each iteration and the Tree-structured Parzen Estimator (TPE) [6] to model the loss. We use the Hyperopt implementation of TPE [7]. Note that this black box optimization method is model-agnostic and applies to any zeroth order optimization problem.

### 4.2 Estimation of Forecast Uncertainty

We estimate parameter uncertainty by approximating the posterior distribution of the parameters given the data. Given a dataset *X* and loss function ℒ (*θ*) as defined in Equation 1, we assume that the likelihood function is tuned to the loss function as

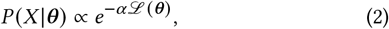

where *α* is a concentration parameter to be estimated as described below. When the loss function ℒ (·) is a regular Bregman divergence, the distribution corresponds to a uniquely determined exponential family [3, 5]. Choosing the parameter prior *P* (*θ*) to be from the conjugate prior family of the resulting exponential family yields a posterior distribution of the form

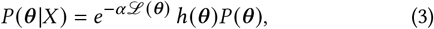

where *h*(*θ*) is uniquely determined by the loss function [14].

Expected values under the posterior distribution can be computed by first sampling from the prior *P θ* with appropriate reweighting and then computing importance sampling estimates. Thus, given *n* parameter samples 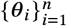 from the reweighted prior, and the corresponding loss function value ℒ(*θ*_*i*_) for each sample, the expected value of any function *g*(*θ*) is estimated as

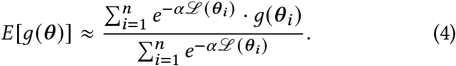

#### Approximate Bayesian Model Averaging

A limitation of this approach is that importance sampling with a broad prior as proposal distribution may require a prohibitively large number of samples. We therefore considered an approximation, hereafter referred to as Approximate Bayesian Model Averaging (ABMA), where we apply Equation 4 not to samples from the prior, but rather to the sequential samples obtained through SMBO. The intuition behind doing so is that these samples represent regions close to the optimal parameter values where the loss is likely to be small, and hence should capture the dominant behavior in sample averages like Equation 4. Additionally, the SMBO implementation is computationally efficient. In Section 5, we show via comparison with an MCMC approach that this is a reasonable approximation. The term *ensemble mean*, used throughout this paper in the context of parameter and forecast averaging, refers to taking the sample means according to Equation 4.

In the application of ABMA, it remains to fix the values of *α* and *n*. We estimate *α* by minimizing the ensemble mean forecast and the ground truth on a validation time interval. To fix *n*, we just choose a value of *n* large enough such that the mean and variance of the parameter samples, computed through application of Equation 4, converges.

#### MCMC-based Estimation of Parameter Uncertainty

A more rigorous method to sample parameter values is via Markov Chain Monte Carlo (MCMC) sampling from the posterior distribution of the parameters given the data, i.e., P (*θ*| *X*). Assuming an appropriate prior over *θ* and a likelihood function *P*(*X*| *θ, s*), where *s* denotes additional parameters in the likelihood function (analogous to *α* in Equation 2), we may consider the larger problem of sampling from the extended posterior distribution

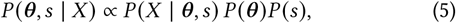

where we assume *θ* and *s* have independent priors. This larger sampling problem is solved by a combination of MCMC and Gibbs sampling steps, using a truncated Normal distribution *Q* (·) (truncated at the boundaries of the search space) as a proposal distribution for MCMC moves. We choose an initial *s*_0_, *θ* _0_, and then employ the following Metropolis-within-Gibbs procedure to alternately sample from the posterior distributions *P* (*θ* |*X, s*) and *P*(*s*| *X, θ*).

1. *MCMC sampling with target distribution P* (*θ* | *X, s*). Given samples *s*_*k*_ −_1_, *θ*_*k*_−_1_ from iteration *k* −1, sample a new *θ*_*k*_ according to the Metropolis criterion, using the proposal distribution *Q* (*θ*_*k*_ | *θ*_*k*_ − _1_).
2. *Sampling s from the posterior distribution P* (*s* | *X, θ*). Given a *θ*_*k*_ sampled in the previous step, sample *s*_*k*_∼ *P* (*s* | *X, θ*_*k*_). This sampling can be made straightforward by choosing a prior for *s* that is conjugate to the likelihood distribution.

The steps above together constitute the Gibbs sampling updates, and are repeated alternately until convergence of the Markov chain. This eventually results in a sample from the extended distribution *P* (*θ, s* | *X*).

#### 4.2.1 Estimating Quantiles for Forecasted Case Count

Given an ensemble of ABMA parameter samples {*θ*_*i*_}, one can compute forecast trajectories for each parameter set in the sample. For a forecast trajectory 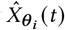, the effective probability density function (pdf) is proportional to 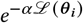. The forecast quantiles on a given day *t* ^*^ are obtained by sorting the forecasted case counts corresponding to different parameter sets on day *t* ^*^ and using the cumulative distribution function (CDF) to find the quantile of interest. This procedure has the desirable property that a trajectory at a fixed quantile value corresponds to a single parameter set. However, forecast trajectories for different parameter sets can cross in time. Therefore quantiles computed at one time point are generally not preserved in time. A work-around is to re-estimate quantiles on each day and piece together a “fixed quantile” trajectory using the forecasted values for that quantile on each day. A trajectory so constructed, however, does not correspond to a single set of epidemiological parameters and is therefore difficult to interpret. We use the former method for the sake of interpretability in our deployment and specify the *t* ^*^ at which quantiles are computed, while the latter method is used for “fixed quantile” forecast comparisons.

### 4.3 Data Spike Smoothing

One of the practical issues we faced was spikes in case count data, originating in delayed reporting of test results and tracking of recoveries and deaths. This leads to accumulated occurrences from the past being reported on a single day. Such spikes therefore need to be smoothed by distributing them several days into the past, even though actual attributed dates were unavailable. We implemented a smoothing technique that carries out this redistribution under some assumptions.

Consider a spike on day *t*_*e*_ in cumulative case counts *X*, given by Δ = *X* [*t*_*e*_] − *X* [*t*_*e*_−_1_] that can be attributed to delayed activity since day *t*_*b*_. We construct a smoothed time series by redistributing Δ as follows:

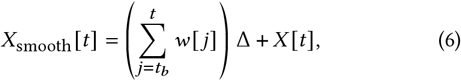

where *t* ∈ {*t*_b_, …, *t* _*e*−1_} and 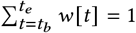. The weights *w* [*t*] can be computed in different ways:

1. Uniform : *w* [*t*] constant.
2. Proportional increments : *w* [*t*] *∝ X* [*t*] − *X* [*t* − 1]
3. Proportional counts: *w* [*t*] *∝ X* [*t*]

Note that a spike in one compartment could correspond to a compensatory dip or a spike in a different compartment such that the total case count stays constant. For example, a spike in recoveries must be compensated by a dip in active cases, since recoveries deplete the active cases. Thus, over the duration of smoothing, the active cases are adjusted by additional recoveries, resulting in a smoothed version of active cases. The model is then fitted on the smoothed case time series rather than on the raw counts.

### 4.4 Epidemiological Model Structure

#### Model Choice Criteria

The choice of epidemiological model was driven by four main criteria: *expressivity*, or the ability to faithfully capture the disease dynamics; *learnability* of parameters conditioned on the available data; *interpretability* in order to understand the evolution of the pandemic; and *generalizability* to future scenarios by incorporating additional information. We observed that simple exponential growth based models that do not account for decreasing susceptibility and finite populations became inaccurate as the pandemic progressed. Agent-based models [17, 38], on the other hand, are overparameterized, with a low level of learnability. While curve-fit models and compartment models had similar levels of accuracy, the parameters of the latter were readily interpretable;^3^ compartmental models, therefore, represented a natural choice.

#### SEIARD Model

We chose a simple, minimal complexity compartmental model that only included compartments for primary case counts or those essential for expressivity. Accessible case count data typically comprised cumulative counts of confirmed cases (*C*), active cases (*A*), recovered cases (*R*), and deceased cases (*D*) with *C* = *A* + *R* + *D*. We assumed confirmed cases to be post-infectious owing to strict quarantining measures. We explicitly modeled the latent “incubation” and “infectious” stages that occur prior to detection so as to allow what-if scenario modeling with changes in isolation and testing policies. Figure 2 depicts the structure of this SEIARD model (see Supplement for the equations). The population is split into compartments S, E, I, A_recov_, A_fatal_, R, D. The states S (Susceptible), E (Exposed), and I (Infectious) and the associated parameters (transmission rate *β*, incubation period *τ*_inc_ = *σ*^−1^, and the infectious period *τ*_inf_ = *γ*^−1^) are defined as in the classic SEIR model [22]. An individual who tests positive moves to a post-infectious but active phase, which is split into A_fatal_ and A_recov_ based on the eventual outcome: fatality (D) with probability *P*_fatal_ or recovery (R). *τ*_fatal_ and *τ*_recov_ are the mean durations from detection to eventual death or recovery respectively. Observed case count data is mapped to compartment populations as follows: *R* = | R |, *D* = |D|, *A* = |A_recov_| + |A_fatal_|.

**Figure 2:**
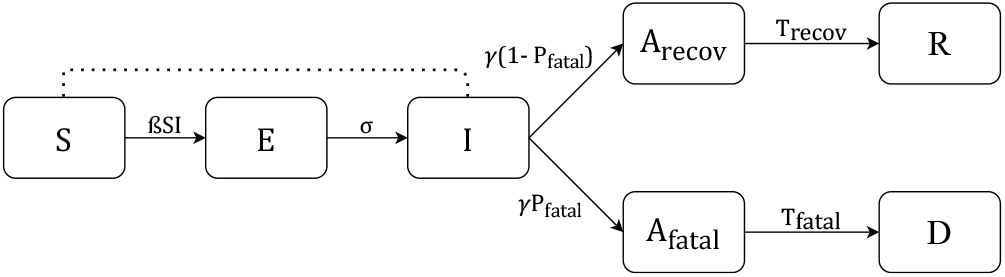
SEIARD Model Structure. Individuals transition between states of disease progression: Susceptible (S), Exposed (E), Infectious (I), Recovered (R), and Deceased (D). Active cases are split between A_recov_ and A_fatal_. Details in Section 8.3.

Interpretable parameters such as reproduction number ℛ _0_ = *β*/*γ, τ*_recov_, and *τ*_fatal_ provid an understanding of the pandemic evolution and reporting gaps, and allow scenario-conditioned forecasting. Changes in the recovery policy and upcoming events such as festivals can be simulated by appropriate adjustment of *τ*_recov_ and *β*.

#### Parameter Fitting Considerations

Estimating the parameters of this model from case counts posed challenges: **a)** Parameter drift due to changes in social distancing policy necessitated refitting the model to a recent time window for every forecast round, **b)** Initial values of the latent compartments^4^ E and I could not be set to 0 because the fitting time interval could occur at an intermediate stage. We treat these latent variables as additional parameters to be optimized during model fitting. More precisely, we consider the ratios of these variables with respect to the initial active counts (at start of fitting interval [*t*_*i*_, *t* _*j*_]) as latent parameters: *E*_active_ratio_ = *E* [*t*_*i*_] /*A* [*t*_*i*_] and *I*_active_ratio_ = *I* [*t*_*i*_] /*A* [*t*_*i*_] **c)** While undetected cases are not explicitly modeled, they are partially accounted for by letting *E* [*t*_*i*_] and *I* [*t*_*i*_] to be free parameters whose fitted values adjust to accommodate the effect of undetected cases. **d)** Identifiability of all key parameters was addressed by incorporating existing domain knowledge about *T*_inc_,*T*_inf_, and *P*_fatal_ from raw (possibly incomplete) case line lists, when available (Figure 3).

**Figure 3:**
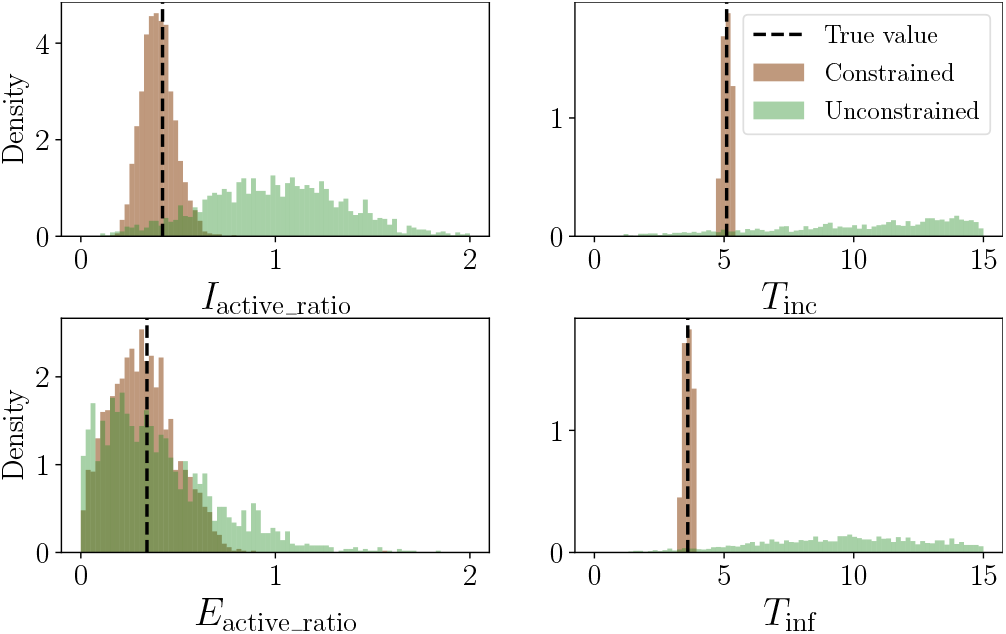
Identifiability of parameters in the SEIARD model. Latent ratio posterior distributions when fitted on synthetic data generated by the true parameter values shown. Distributions were estimated using the MCMC method of Section 4.2 under a broad prior as well as under fixed choices of *τ*_inc_, *τ*_inf_, and *P*_fatal_.

## 5 EXPERIMENTAL RESULTS

To demonstrate the value of the proposed framework, we report (a) empirical results validating choices of hyperparameters, uncertainty estimation, and data preprocessing, (b) field performance and impact of the deployed system in Mumbai, and (c) empirical comparison of our approach with other state-of-the-art (SOTA) models in Reich Lab on COVID-19 case data from the US. The main elements of our experiment setup are:

### Data

We consider time series of confirmed (*C*), active (*A*), reported (*R*), and deceased (*D*) cases. Results shown are based on real data from the city of Mumbai except for the hyperparameter optimization discussion (Covid19India data) and the comparison with Reich Lab models (JHU data on 45 US regions).

### Algorithms

Various alternatives within our proposed framework (ABMA vs. MCMC, smoothing variants) as well as 26 different models from Reich Lab were considered.

### Evaluation Metric

Mean Absolute Percentage Error (MAPE) along each of the four variables (*C, A, R, D*) and aggregated over the compartments with weights is used as the primary metric for evaluation and comparisons. Uncertainty estimation is discussed in terms of credible intervals.

### 5.1 Data Spike Smoothing

Our experiments indicated that smoothing (in particular, the Proportional count variant) leads to better prediction of future cases. We evaluated the smoothing variants discussed in Section 4.3 on simulated incident (new) raw data *X*_inc_[*t*] with a simulated spike caused due to delays in data reporting between dates *t*_*b*_ and *t*_*e*_. Following on-the-ground reporting discrepancies, we assume that only a fraction *δ*_*t*_ of the true incident cases *X*_inc_[*t*] on day *t* are actually reported on that day, i.e., *X* [*t*] = *X* [*t* − 1] + *δ*_*t*_*X*_inc_ [*t*]. The remaining (1 − *δ*_*t*_)*X*_inc_ [*t*] cases are reported on the end date *t*_*e*_, resultin g in a case spike on day *t*_*e*_, i.e., 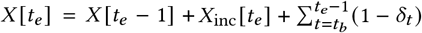 · *X*_inc_ [*t*]. For the simulation, we assume that the *δ*_*t*_ are i.i.d. samples from a Beta distribution (here, chosen as Beta 2, 2). Other parameters are chosen according to Table 4. Smoothing methods are then applied to the simulated data generated after the dependent case counts are adjusted to sum to the confirmed cases.

Table 1 and Figure 4 show the results of smoothing experiments. Overall, we found that the “Proportional counts” smoothing method works best at reconstructing the raw data with reasonable accuracy.

**Table 1:**
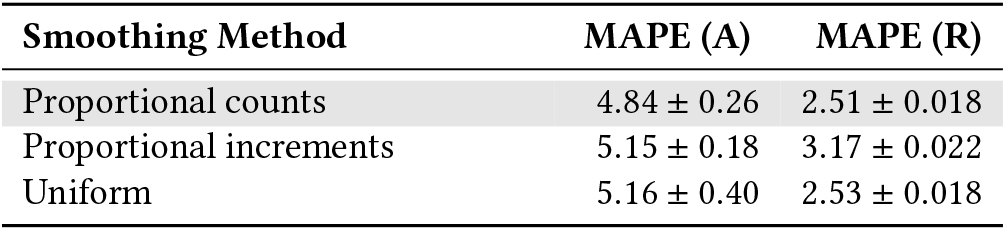
Comparison of smoothing methods on simulated spiky data. Spikes were generated as described in the text on 5000 simulated time series, generated via uniform sampling of parameter values in Table 4 (a, b) (Supplement). MAPE loss on Active (*A*) and Recovered (*R*) compartments was computed between the smoothed and the true, unspiked series.

| Smoothing Method | MAPE (A) | MAPE (R) |
| --- | --- | --- |
| Proportional counts | $4.84 \pm 0.26$ | $2.51 \pm 0.018$ |
| Proportional increments | $5.15 \pm 0.18$ | $3.17 \pm 0.022$ |
| Uniform | $5.16 \pm 0.40$ | $2.53 \pm 0.018$ |

**Figure 4:**
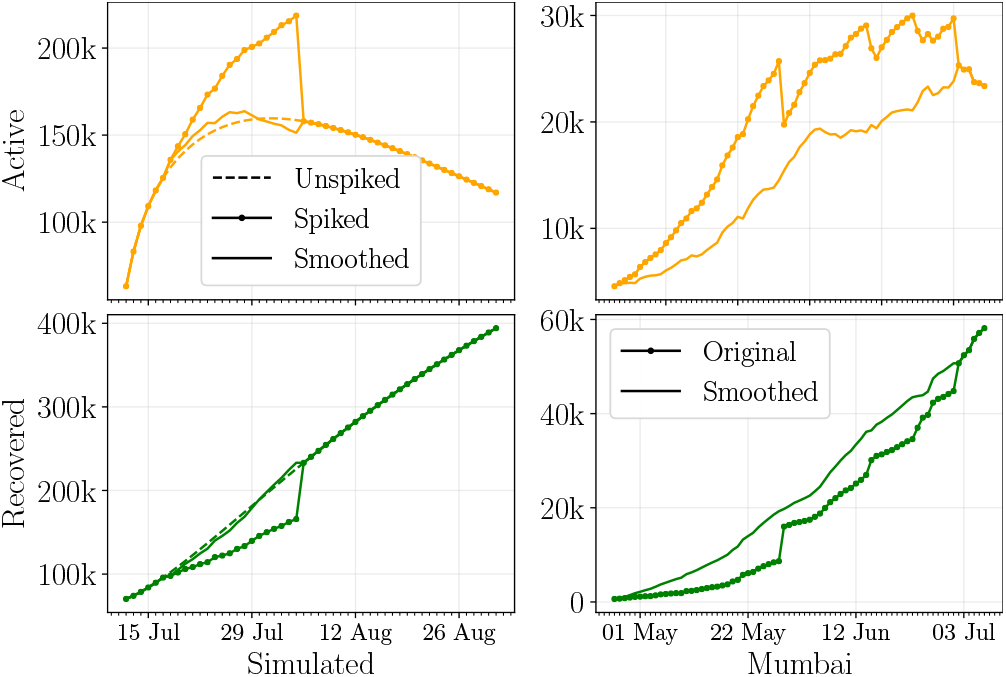
Performance of smoothing algorithm on simulated spike and Mumbai data. Smoothing is done via the “Proportional counts” method.

### 5.2 Hyperparameter Optimization

Within our modeling framework, the following hyperparameters need to be tuned to achieve good performance: a) *T*_*θ*_, the length of the fitting period in days to estimate model parameters *θ*, b) *T*_*α*_, the number of days of data used to tune the concentration parameter *α*, and c) *n*, the number of trials used to optimise *θ*. In addition, it was also necessary to identify a suitable loss function and compare the performance of the TPE to other fitting techniques. Details of these tuning experiments are given in the Supplement.

### 5.3 Parameter and Uncertainty Estimation: ABMA vs. MCMC

We compared the ABMA method of Section 4.2 to the MCMC sampling approach described there, along three dimensions: a) comparison of the lowest loss values achieved, b) comparison of case count forecasts, and c) distributional comparison of the sampled parameters. For these comparison experiments we chose a duration of 28 days for fitting, from 1 October to 28 October 2020, and a duration of 21 days, from 1 November to 21 November 2020, for evaluation. The three days in between were used for fitting *α* in the ABMA procedure. The time taken to generate MCMC samples was noticeably higher than that for ABMA: under a controlled compute setting, ABMA was found to be an order of magnitude faster. This was one of the principal motivations behind the choice of ABMA. Details of MCMC sampling as well as comparison of sampled parameters are given in the Supplement.

#### Comparison of the best loss values achieved

We evaluated the MAPE losses across forecasted ensemble mean time series 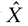 on the test duration, corresponding to parameters sampled by ABMA vs. MCMC algorithms (Figure 5). Note that for MCMC samples, the ensemble mean forecasts are generated by simply averaging the forecasts for the sampled parameter sets, while for ABMA samples, the ensemble mean forecasts are generated by weighting each forecast by its importance sampling factor. The figure illustrates that the differences between the MAPE loss values are minor when comparing both ensemble means on the test set.

**Figure 5:**
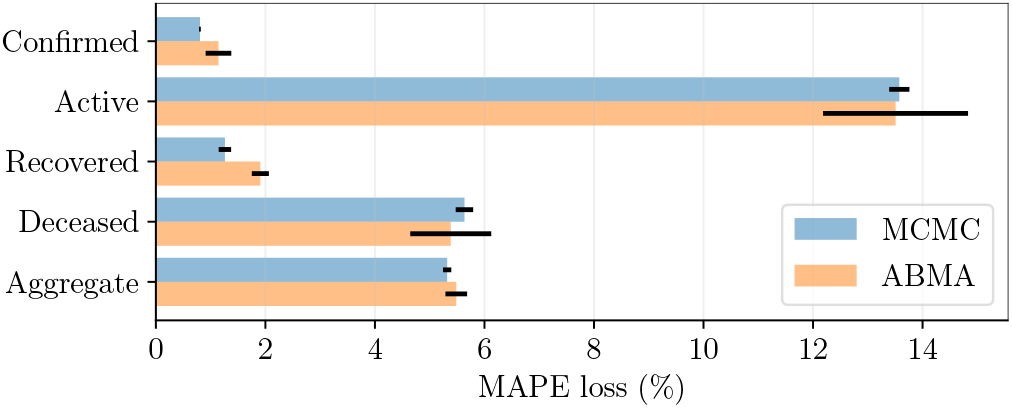
Comparison of test losses for MCMC and ABMA. MAPE losses for the ensemble mean forecast on the test period are shown for each compartment under the two methods. Error bars denote the standard error across 10 runs.

#### Comparison of case count forecasts

We now turn to a comparison of the forecasts themselves. Figure 6 shows comparisons of ensemble mean forecasted values, and their 95% credible intervals for the four compartments studied which were generated using quantiles recomputed daily. We find that mean values and intervals agree quite well for all compartments except active cases. Also, for active cases, interestingly the true case count is observed to fluctuate between the MCMC and ABMA values.

**Figure 6:**
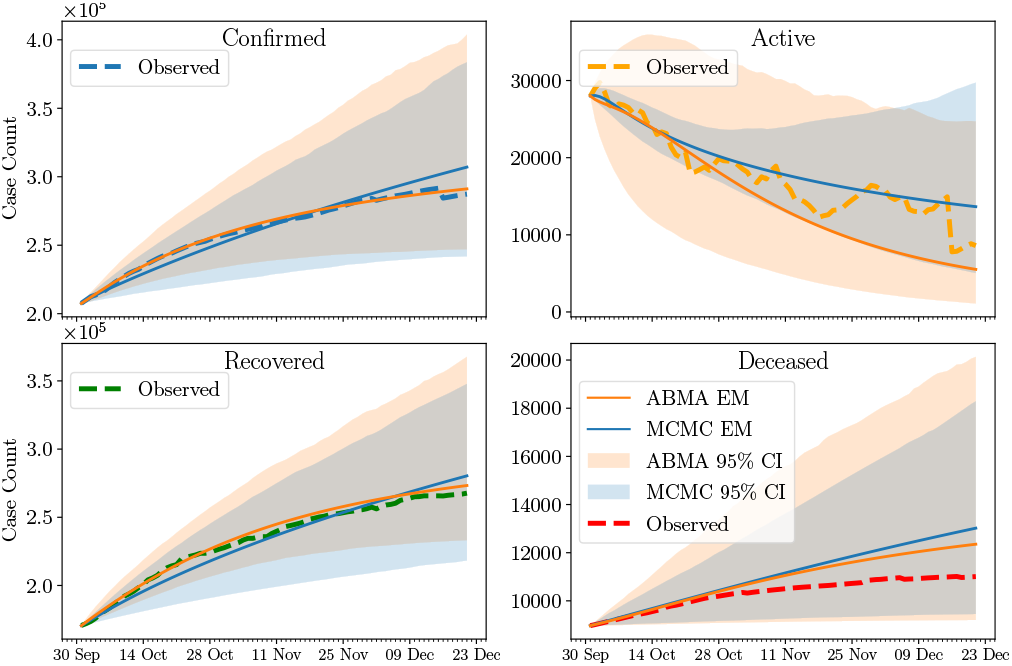
Comparison of 95% Credible Intervals and Ensemble Mean of MCMC and ABMA. ‘EM’ denotes the Ensemble Mean forecast. Credible intervals (CI) are shown along with the point forecasts.

### 5.4 Field Deployment Performance

The ABMA framework was used to provide actionable insights for the city of Mumbai, India from May 2020 to October 2020. It is also currently being used to forecast hospital burden in the state of Jharkhand, India. Figure 7, showing the ensemble mean forecast against the ground truth, and Table 2, showing the estimated parameters and loss values, confirm the high accuracy and interpretability of our methodology across different phases of the first wave of the pandemic.

**Figure 7:**
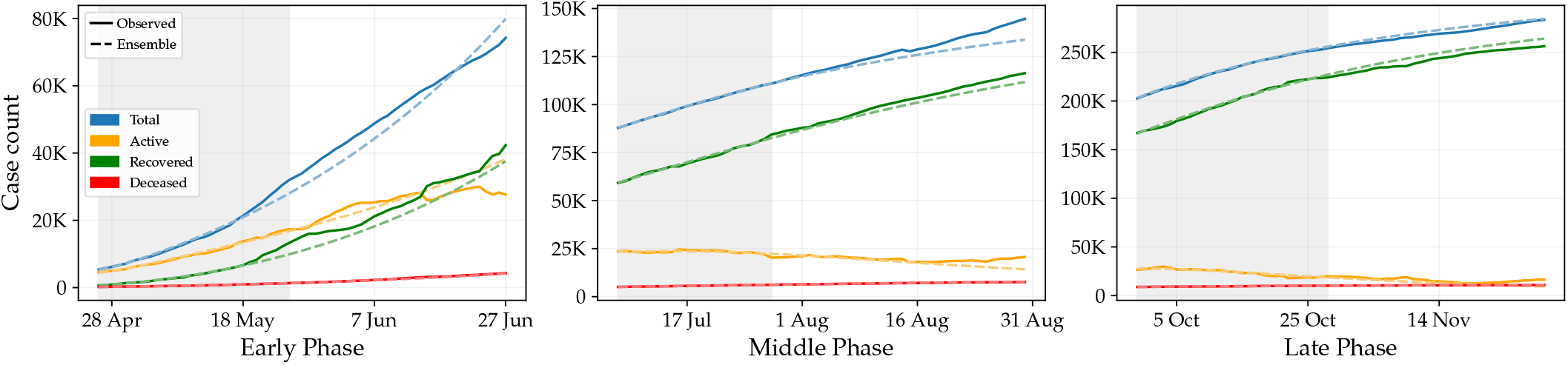
Performance of ABMA on Predicting Mumbai Caseload. Predicted and ground truth case counts for Mumbai city across compartments and phases of the pandemic. Here, the parameter fitting period is *T*_*θ*_ = 30 days and the fitting period for the hyperparameter *α* is *T*_*α*_ = 3 days (see Supplement). Forecasts are shown 30 days beyond the end of the *T*_*α*_ fitting period. All dates refer to the year 2020.

**Table 2:**
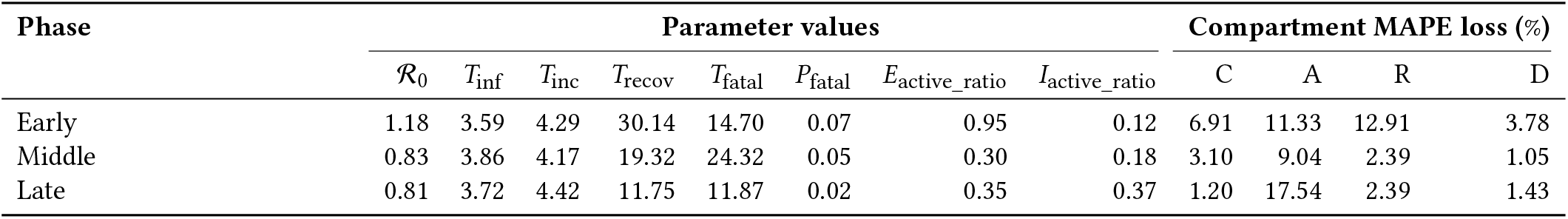
Quantitative Performance of ABMA on Mumbai. Ensemble mean (ABMA) *parameters*, and test MAPE loss (%) for each *compartment* of the ABMA forecasts. Note that the ABMA forecast is the mean forecast, not the forecast of the mean parameters.

Recommendations made using this forecasting framework helped increase Mumbai’s ICU bed capacity by over 1200 units, with 95% utilization of ICUs at when hospitalizations peaked. Moreover, over the deployment period, no absolute shortfall of critical health care resources became apparent.

### 5.5 Comparison with SOTA Models

We used the ReichLab forecasting hub [1] for US states as a basis for comparing the performance of our forecasting framework with other methods (Table 3). The source of data for the forecasts is the JHU CSSE data [18]. We evaluated our method only on regions for which all four primary case counts were available (44 states plus Washington D.C.). Parameter and hyperparameter choices for the evaluation are in the Supplement (Table 4). The basis for comparison with other models is the MAPE value on deceased case counts.

**Table 3:** Performance of top 10 Reichlab models submitting forecasts for at least 45 regions on cumulative death counts. All models were fitted on data from 18 Aug to 19 Sept 2020. Based on hyperparameter optimisation for Mumbai, we chose *T*_*θ*_ = 30, *T*_*α*_ = 3. Model forecasts four weeks into the future starting 20 Sept 2020, aggregated every week, were evaluated by computing their MAPE values for every region, and then taking the median value across all regions (reported here).

| Rank | Model | Median MAPE (%) |
| --- | --- | --- |
| 1 | UMass-MechBayes | 1.21 |
| 2 | Karlen-pypm | 1.32 |
| 3 | SteveMcConnell-CovidComplete | 1.33 |
| 4 | **ABMA** | **1.38** |
| 5 | YYG-ParamSearch | 1.44 |
| 6 | UCLA-SuEIR | 1.49 |
| 7 | PSI-DRAFT | 1.65 |
| 8 | DDS-NBDS | 1.70 |
| 9 | CEID-Walk | 1.77 |
| 10 | COVIDhub-baseline | 1.81 |

**Table 4:**
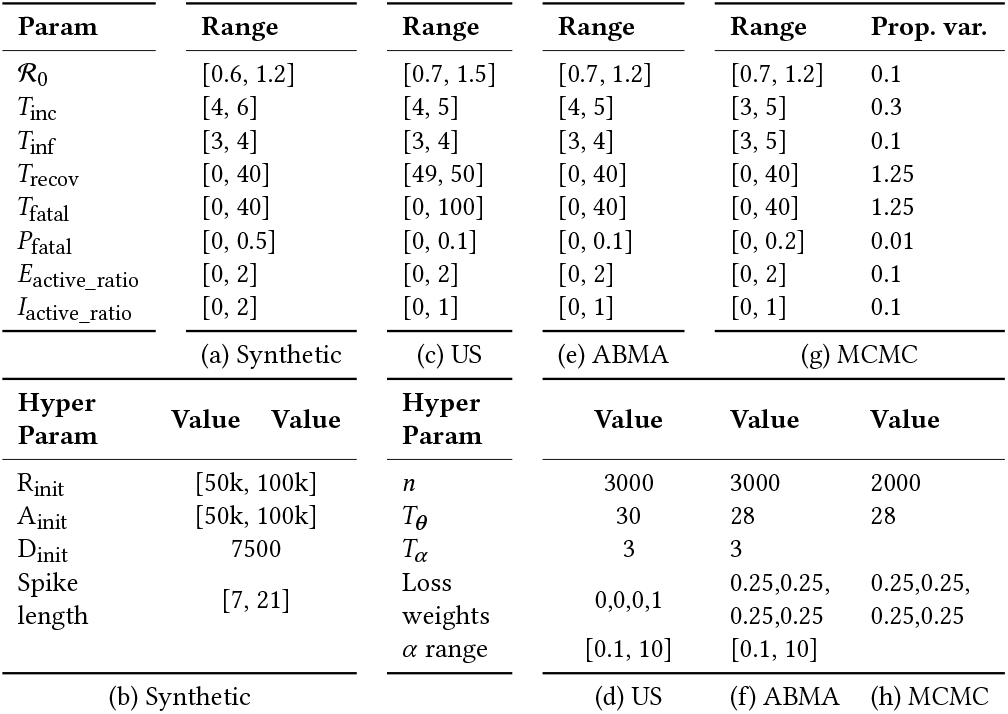
Parameter and hyperparameter ranges for all experiments. (a-b) simulated smoothing data and spikes; (c-d) ReichLab comparison experiments; (e-f) ABMA approach applied to Mumbai data; (g-h) MCMC approach applied to Mumbai data. Subscript “init” subscript refers to the corresponding initial value. The four loss weight values are for the four series C, A, R, D respectively.

We found that 26 models submitted to ReichLab had submissions for at least 45 regions over the duration studied, with a range of median MAPE values between 1.21% and 4.26%. The top 10 models by median MAPE are shown in Table 3. Our forecasts have low error and compare well to other models without the need for any region-specific customisation of hyperparameters or fitting method. We further analyzed the distribution of errors across regions (Figure 8) and assessed performance relative to other modeling approaches for each US region using normalized MAPE defined as

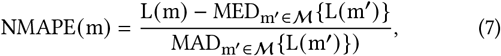

where L (*m*) is the MAPE loss for model *m*, ℳ is the set of all models, MED and MAD denote the Median and the Median Absolute Deviation operators respectively. The variation in relative model performance across states could be attributed to changes in social distancing policies and the consequent variations in disease spread dynamics.

**Figure 8:**
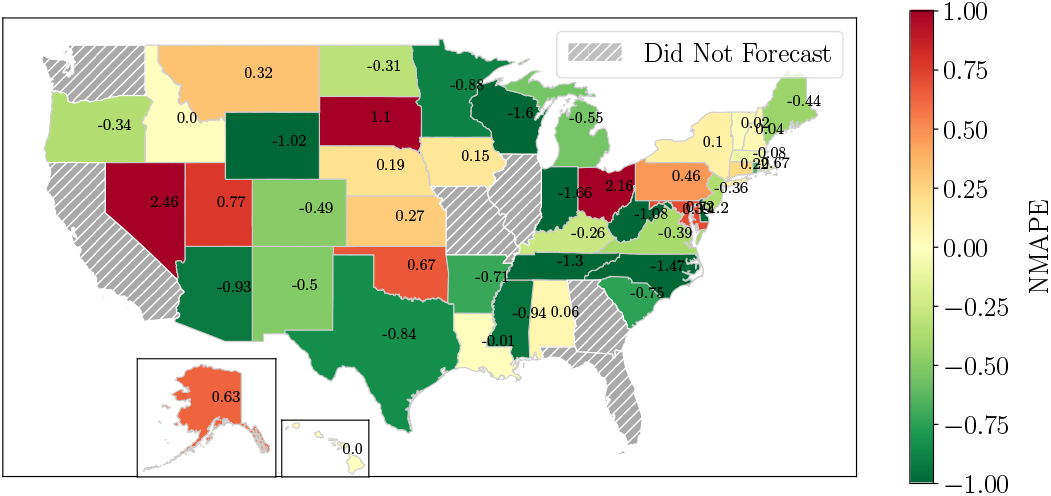
Model Performance on US States. NMAPE is the normalized MAPE (green values correspond tolower error) defined in the text (Equation 7).

## 6 LESSONS LEARNED

### Dynamic data entails human in the loop

The data-related challenges mentioned in the Introduction led to us relying heavily on humans in the loop for tracking evolving data definitions and carrying out semi-automated quality checks. Data versioning and pre-processing prior to modeling (Section 5.1) were also essential.

### Model interpretability and identifiability is paramount

Our choice of the SEIARD model (Section 4.4) over other model families was motivated by planning needs and policy choices. Therefore the parameters had to be interpretable, independently verifiable where possible, and robustly estimable from the available data.

### Application needs should dictate modeling choices

Focus on capacity planning had a tangible meaning for the horizon of interest: policies around capacity took about a month to implement. We customized our model fitting with loss computed over this time horizon. When provided with information on upcoming policy changes and events—festivals, for example—we adapted the models accordingly to generate accurate forecasts.

### Insights must be actionable

Model insights had to be translated to concrete action guidance to enable smooth planning. Uncertainty estimation (Section 4.2) allowed us to provide three relevant scenarios: a) a planning b) a high case, c) a low case scenario. Region-specific testing levels, evolving severity of cases and sero-surveillance information to understand the state of the pandemic were important factors informing the selection of planning scenarios.

### Capacity gaps at the last mile are hard to anticipate

Although there were no apparent capacity shortfalls at a city level, the ability of a critical patient to access these resources is mediated by granular factors such as access to information on the availability of beds, local emergency transportation, ability to pay, and other equity considerations, which need to be factored in.

## 7 CONCLUSIONS AND FUTURE DIRECTIONS

We have presented a flexible modeling framework and demonstrated its value for epidemic forecasting using the kind of case count aggregate data that is typically available in a constrained public health setting. The deployed system was used to drive decisionmaking and planning with good accuracy (worst case MAPE *<* 20%) during the COVID-19 pandemic in Mumbai and Jharkhand, India. Our framework enables rapid forecasting with uncertainty estimates, and is extensible to other model families and to different loss functions. It also enables the optimization of hyperparameters such as fitting durations and ensemble weights. We motivated the choice of the specific compartmental model used via identifiability of the underlying parameters given the data constraints. Empirical comparison of our methods with other advanced models in the ReichLab hub on real-world data further points to their efficacy.

In future work, we plan to open-source this framework, create a playbook around it, and apply it to the estimation of case burden in other infectious diseases, such as Tuberculosis, which are widespread across the developing world. One of the limitations of the current framework that we recognize is that the underlying parameters are static. We plan to extend this modeling approach to capture time-varying parameters that are yet interpretable, and thus enable better forecasting.

## Data Availability

The majority of the data used in this study was provided to us by Brihanmumbai Municipal Corporation (BMC), with whom we had a partnership. This data was also publically available on www.covid19india.org.
The rest of the data used for this study was publically available on https://github.com/CSSEGISandData/COVID-19.

## Acknowledgements

We would like to thank Janak Shah and the WIAI Programs team for their support with data and deployment processes. This study is made possible by the generous support of the American People through the United States Agency for International Development (USAID). The work described in this article was implemented under the TRACETB Project, managed by WIAI under the terms of Cooperative Agreement Number 72038620CA00006. The contents of this manuscript are the sole responsibility of the authors and do not necessarily reflect the views of USAID or the United States Government. This work is co-funded by the Bill and Melinda Gates Foundation, Fondation Botnar and CSIR - Institute of Genomics and Integrative Biology. It was enabled by several partners through the informal COVID19 data science consortium, most notably Shreyas Shetty, Anupama Agarwal, and a group of volunteer data scientists under the umbrella group “Data Science India vs. COVID”. We thank Amazon Web Services for Cloud credits and support, the Smart Cities Mission for providing early impetus to this work, and the Brihanmumbai Municipal Corporation and Integrated Disease Surveillance Program (Jharkhand) for support while facing unprecedented challenges.

## 8 SUPPLEMENT

We list additional details required to reproduce the reported results.

### 8.1 Parameter Searchspace

Table 4 below lists the parameter searchspace and hyperparameter ranges used in the experiments described in the main text.

### 8.2 Hyperparameter Optimisation

Since we wished to assess the robustness of optimal hyperparameters to choice of location, we performed experiments for several regions of India that were highly impacted by the pandemic: Mumbai, Pune, Delhi and Kerala. To facilitate comparisons, we used case count data from public sources [13], across two different stages of the pandemic: our evaluations were carried out on case counts from July 1 to 28, 2020 and November 1 to 28, 2020. Fitting was carried out over durations prior to these dates. The evaluation metric used was the MAPE value on the entire evaluation period.

Due to the prohibitively large search space, a complete grid search over the hyperparameter choices was not feasible. We thus ordered the experiments and performed them sequentially, optimising one or more parameters in each step and fixing them in subsequent experiments to optimise other parameters.

We first performed a grid search over combinations of *T*_*θ*_ and *T*_*α*_ (Figure 9) to find the best choices. We found within the selected fitting period *n* ⪆ 3000 trials were sufficient for convergence of sample mean parameters, and thus chose this value of *n*. The application loss of interest is MAPE, however, in addition to this, we explored two other loss functions during fitting as well as *α* estimation:root mean square error (RMSE) and root mean square logarithm error (RMSLE). The MAPE on the evaluation period with the three different fitting configurations was 5.50 (MAPE), 5.96

**Figure 9:**
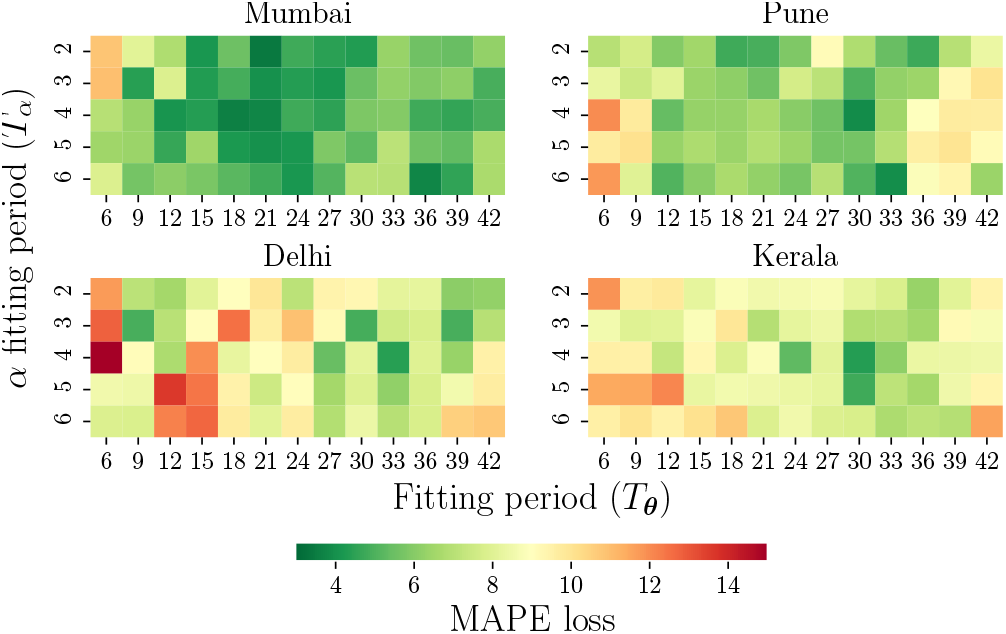
Heatmap of MAPE values on test periods as a function of *T*_*θ*_ and *T*_*α*_. MAPE is computed on a four week evaluation period and averaged across experiments for two time periods. The optimal values of (*T*_*θ*_, *T*_*α*_) are – Mumbai: (21, 2), Pune: (30, 4), Delhi: (33, 4), Kerala: (30, 4), Overall: (30, 3). “Overall” corresponds to the lowest mean MAPE across all four regions.

(RMSE) and 15.81 (RMSLE). Based on the results, the loss function selected was MAPE for both fitting and *α* estimation.

### 8.3 SEIARD Model Dynamics

The dynamical equations governing the transitions in SEIARD model (Figure 2) are

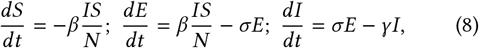

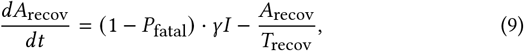

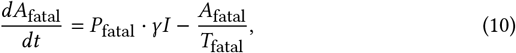

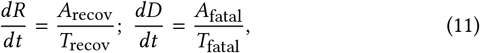

where *N* is the city population, *β, σ*, and *γ* are the standard epidemiological parameters for a SEIR model, *P*_fatal_ is the transition probability to the mortality branch, and *T*_recov_ and *T*_fatal_ are timescales that govern transitions out of the A_recov_ and A_fatal_ compartments. Variables *S, E, I, A*_recov_, *A*_fatal_, *R, D* denote the populations of the similarly named compartments.

### 8.4 MCMC Implementation Details

Let *X*[*t*] = [*X*_*h*_ [*t*]]_*h*∈ℋ_ be a multivariate time series with *X*_*h*_ [*t*] denoting the *h*^th^ compartment time-series, and ℋ be the set of indices of components. Let the fitting period be given by [*t*_*i*_, *t* _*j*_] The key components of our MCMC-within-Gibbs sampling are:

#### Likelihood function

We assume a likelihood function of the form

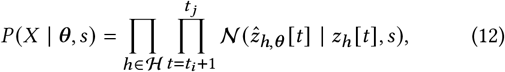

where 𝒩 (*z* | *μ, σ*^2^) denotes the Normal distribution pdf with mean *μ* and variance *σ*^2^ following an appropriate conjugate prior. Further, *z*_*h*_ [*t*] = log(*X*_*h*_ [*t*]) − log(*X*_*h*_ [*t* − 1]), and 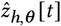 is the forecast equivalent of *z*_*h*_ [*t*].

#### Proposal distribution

At iteration *k*, we generate the samples from the proposal distribution for accept-reject step as

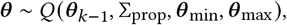

where *θ*_*k*−1_ is the parameter vector chosen at *k*−1, and *Q*(·) is the pdf of a multivariate truncated Gaussian with the parameter range [*θ* _min_, *θ*_max_] and covariance matrix Σ_prop_ as listed in Table 4(g) (columns: Range and Prop. var).

We further assume that *s*, the variance of the normal likelihood, has the conjugate prior *s* ∼ InvGamma (*u, v*). Thus, it is straightforward to show, by multiplying the Normal likelihood with the prior, that if the MCMC chain has sampled parameters *θ*_*k*_, the sample *s*_*k*_∼ *P*(*s*_*k*_| *θ*_*k*_, *X*) is also drawn from an Inverse Gamma distribution with parameters

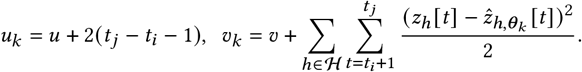

Parameter ranges for our implementation are in Table 4. The MCMC implementation was adapted for *X* = [*C, A, R, D*] with 5 chains of length 25*k*, a stride of 5 for sampling, a burn-in length of 50% of the chain, *u* = 40, and *v* = 2/700.

### 8.5 Parameter distribution: ABMA vs. MCMC

To validate the uncertainty estimates of Section 5.3, we compared the samples collected from both methods aggregated over 10 runs of each method (Figure 10). Major distributional differences are found only in ℛ_0_ and *E*_active_ratio_, but even for these parameters there is significant overlap between the distributions sampled by the two methods. Note that distributions for the ABMA approach are weighted by the importance sampling factor exp(−*α*ℒ(*θ*)).

**Figure 10:**
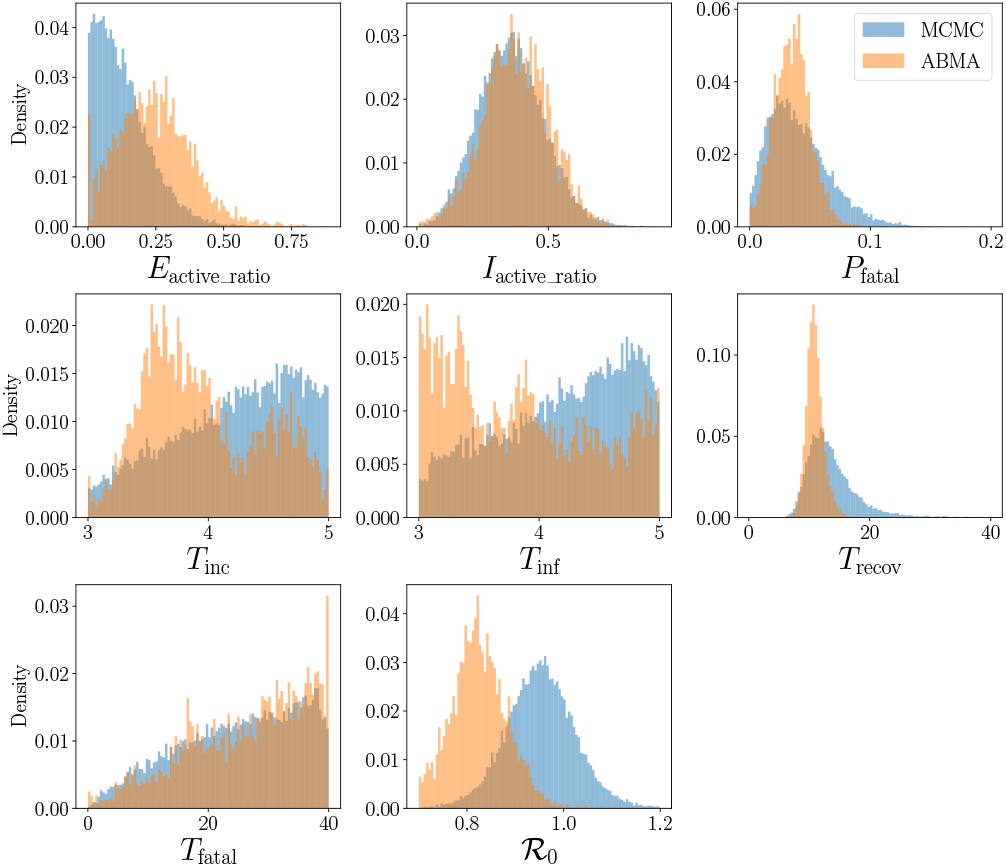
Empirical distributions of ABMA and MCMC samples. We observe large distributional overlap for all parameters.

### 8.6 Uncertainty Evaluation

To compare the efficacy of our uncertainty estimation, we compared the forecast distributions of the models submitted to ReichLab (Section 5.5) using the quantile loss described below:

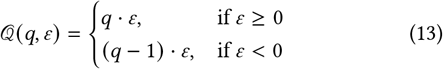

where *ε* is the error between the forecasted quantile *q* and the ground truth. In our case, the error is measured by the MAPE loss function. For every (model, quantile) tuple, we computed a median quantile loss value aggregated across regions. ABMA quantiles are generated by recomputing them daily. Figure 11 shows the variation of the median quantile loss across quantiles for all models. The loss curves of our model and the UMass-MechBayes model, the top performing model according to Table 3 are highlighted. While our model is one of the best around the 50^*th*^ percentile, it exhibits higher relative loss at extreme quantiles.

**Figure 11:**
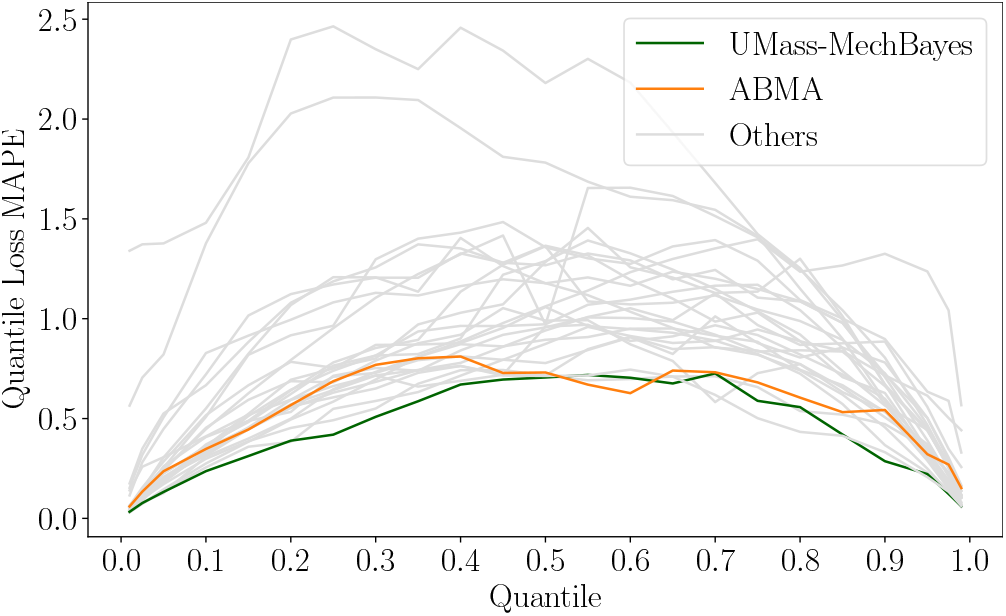
Median Quantile MAPE Loss for 26 models from ReichLab. Median is taken across all states. Our model and the top ranked model in Table 3 are marked.

### 8.7 Report Template

**Figure 12:**
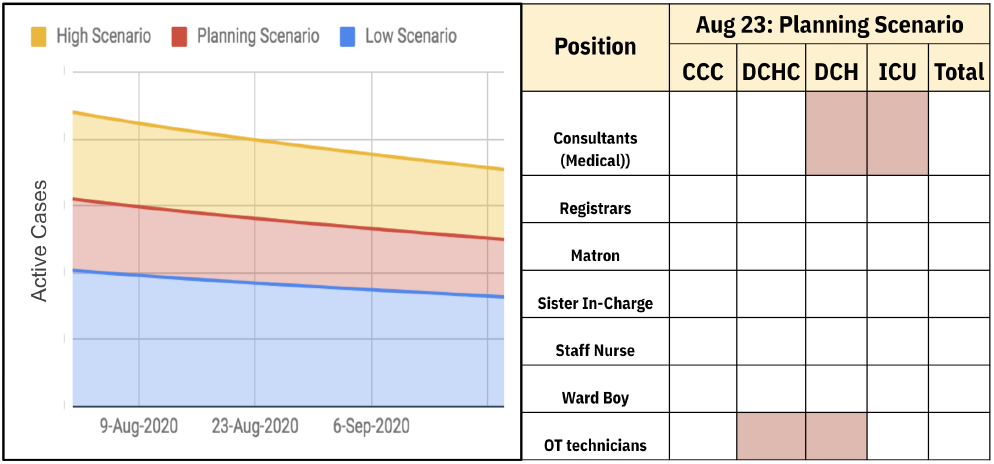
Sample report template shared with city officials: Personnel recommendations and shortfall (red boxes) for various facilities. The facilities are CCC (Covid Care Center), DCH (Dedicated Covid Hospital), DCHC (Dedicated Covid Health Center), ICU (Intensive Care Unit).

## Footnotes

1 Within the Amazon Web Service framework: Lambda functions for ingesting and cleaning, Glue for collating, Athena for exposing.

2 Publishers log to ML tracking platforms such as MLFlow and Weights & Biases.

3 Application of compartmental models to a few Indian districts led to estimates of recovery period exceeding 50 days, leading to the discovery of a serious data reporting gap. Interpretability of these parameters, therefore, turned out to be important.

4 *S* is not viewed as latent since the net population can be assumed to be nearly constant.

## REFERENCES

[1] Reichlab UMass Amherst. 2020. COVID-19 Forecast Hub. https://github.com/reichlab/covid19-forecast-hub

[2] F. Balabdaoui and D. Mohr. 2020. Age-stratified model of the COVID-19 epidemic to analyze the impact of relaxing lockdown measures: nowcasting and forecasting for Switzerland. medRxiv (2020).

[3] Banerjee et al. 2005. Clustering with Bregman Divergences. J Mach Learn Res 6, 58 (2005), 1705–1749.

[4] N. Bannur et al. 2020. Synthetic Data Generation for Improved COVID-19 Epidemic Forecasting. medRxiv (2020).

[5] O. Barndorff-Nielsen. 1978. Information and Exponential Families in Statistical Theory. Wiley.

[6] J. Bergstra et al. 2011. Algorithms for Hyper-Parameter Optimization. In NeurIPS. 2546–2554.

[7] J. Bergstra et al. 2013. Making a science of model search: Hyperparameter optimization in hundreds of dimensions for vision architectures. In Intl. conf. on machine learning. 115–123.

[8] Bertozzi et al. 2020. The challenges of modeling and forecasting the spread of COVID-19. PNAS 117, 29 (2020), 16732–16738.

[9] L. Bettencourt and R. Ribeiro. 2008. Real Time Bayesian Estimation of the Epidemic Potential of Emerging Infectious Diseases. PLOS ONE 3, 5 (May 2008), 1–9.

[10] Bhardwaj et al. 2020. Robust Lock-Down Optimization for COVID-19 Policy Guidance. In AAAI Fall Symposium.

[11] Capaldi et al. 2012. Parameter estimation and uncertainty quantification for an epidemic model. Mathematical biosciences and engineering 9 (Jul 2012), 553–76.

[12] G. Chowell. 2017. Fitting dynamic models to epidemic outbreaks with quantified uncertainty: A primer for parameter uncertainty, identifiability, and forecasts. Infectious Disease Modelling 2, 3 (2017), 379–398.

[13] Covid19India. 2020. Coronavirus in India: Latest Map and Case Count. https://www.covid19india.org/

[14] P. Diaconis and D. Ylvisaker. 1979. Conjugate Priors for Exponential Families. Ann. Statist. 7, 2 (Mar 1979), 269–281.

[15] Elderd et al. 2006. Uncertainty in predictions of disease spread and public health responses to bioterrorism and emerging diseases. PNAS 103, 42 (2006), 15693–15697.

[16] Y. Fang et al. 2020. Transmission dynamics of the COVID-19 outbreak and effectiveness of government interventions: A data-driven analysis. Journal of Medical Virology 92, 6 (2020), 645–659.

[17] Ferguson et al. 2020. Report 9: Impact of non-pharmaceutical interventions (NPIs) to reduce COVID-19 mortality and healthcare demand.

[18] Center for Systems Science and Engineering at Johns Hopkins University. 2020. COVID-19 Data Repository. https://github.com/CSSEGISandData/COVID-19/

[19] S. Funk et al. 2019. Assessing the performance of real-time epidemic forecasts: A case study of Ebola in the Western Area region of Sierra Leone, 2014-15. PLoS computational biology 15, 2 (2019), e1006785.

[20] G. Giordano et al. 2020. Modelling the COVID-19 epidemic and implementation of population-wide interventions in Italy. Nature Medicine 26 (Jun 2020), 1–6.

[21] IHME COVID-19 health service utilization forecasting team. 2020. Forecasting the impact of the first wave of the COVID-19 pandemic on hospital demand and deaths for the USA and European Economic Area countries. medRxiv (2020).

[22] H. Hethcote. 2000. The Mathematics of Infectious Diseases. SIAM Rev. 42, 4 (Dec 2000), 599–653.

[23] Jones. 2001. A taxonomy of global optimization methods based on response surfaces. J. of global optimization 21, 4 (2001), 345–383.

[24] Kaggle. 2020. COVID19 Global Forecasting. https://www.kaggle.com/c/covid19-global-forecasting-week-1

[25] Kermack and A. McKendrick. 1927. A contribution to the mathematical theory of epidemics. Proc. R. Soc. Lond. A 115, 4 (1927), 700–721.

[26] S. Mandal et al. 2020. Prudent public health intervention strategies to control the coronavirus disease 2019 transmission in India: A mathematical model-based approach. Indian Journal of Medical Research 151, 2 (2020), 190–199.

[27] G. Massonis et al. 2020. Structural identifiability and observability of compart-mental models of the COVID-19 pandemic. Annu Rev Control (Dec 2020).

[28] S. Moghadas et al. 2020. Projecting hospital utilization during the COVID-19 outbreaks in the United States. PNAS 117 (Apr 2020), 9122–9126.

[29] Covid Act Now. 2020. America’s COVID warning system. https://covidactnow.org

[30] M. Paggi. 2020. Simulation of Covid-19 epidemic evolution: are compartmental models really predictive? 2004.08207

[31] T. Philipson. 2000. Chapter 33 Economic epidemiology and infectious diseases. Handbook of Health Economics, Vol. 1. Elsevier, 1761–1799.

[32] K. Roosa and G. Chowell. 2019. Assessing parameter identifiability in compart-mental dynamic models using a computational approach: application to infectious disease transmission models. Theor. Biol. Medical Model. 16 (Jan 2019).

[33] P. Samuia et al. 2020. A mathematical model for COVID-19 transmission dynamics with a case study of India. Chaos, Solitons & Fractals 140 (2020), 110173.

[34] M. Serhani et al. 2020. Mathematical modeling of COVID-19 spreading with asymptomatic infected and interacting peoples. J. Appl. Math. Comput. 17 (Aug 2020), 1–20.

[35] F. Tabataba et al. 2017. A Framework for Evaluating Epidemic Forecasts. BMC Infectious Diseases 17 (May 2017), 345.

[36] N. Thakkar et al. 2020. Social distancing and mobility reductions have reduced COVID-19 transmission in King County, WA. Institute for Disease Modeling.

[37] P. Walker et al. 2020. The impact of COVID-19 and strategies for mitigation and suppression in low-and middle-income countries. Science 369, 6502 (2020), 413–422.

[38] B. Wilder et al. 2020. Modeling between-population variation in COVID-19 dynamics in Hubei, Lombardy, and New York City. PNAS 117, 41 (2020), 25904–25910.

